# Dynamic balance recovery in chronic Acquired Brain Injury participants following a perturbation training

**DOI:** 10.1101/2021.01.25.21250435

**Authors:** Katherin Joubran, Simona Bar-Haim, Lior Shmuelof

**Author notes:** **Corresponding authors**: Katherin Joubran, PhD, Nazareth, P.O.Box 2819, Israel 16503., Lior Shmuelof, PhD, Ben-Gurion University of the Negev, Beer-Sheva, 84105, Israel. **Conflicts of interest**: Dr Simona Bar-Haim holds equity positions in Step of Mind, the company that developed the Re-Step system. **Clinical trial registration number**: the trail is registered at the ClinicalTrails.gov, NCT02215590.

## Abstract

**Background:** Acquired Brain Injury (ABI) is defined as a damage to the brain that occurs after birth. Subjects post-ABI suffer from dynamic balance impairments that persist years after the injury.

**Objective:** To explore the effect of a perturbation method which is consisted of unexpected balance perturbations using Re-Step™ technology on the recovery of dynamic balance and gait velocity in chronic ABI participants.

**Methods:** In a clinical trial, 35 chronic ABI participants (stroke and traumatic brain injury) participated in 22 sessions of perturbation-training, twice a week for 3 months. Dynamic balance was assessed pre and post-training using Community Balance and Mobility Scale (CB&M). Gait velocity was also assessed in the stroke participants using the 10-meter walk test (10MWT).

**Results:** Dynamic balance improved significantly post-training (*p*=0.001). This improvement was greater than the improvement that was observed in a sub-group that was tested twice before training (*p*=0.04). 16 participants (45.7%) out of 35 met or exceeded minimal detectable change (MDC) of the CB&M Scale. Self-paced velocity also improved significantly (*p*=0.02) but only 2 participants (9.5%) out of 21 exceeded the MDC of 10MWT post-stroke.

**Conclusions:** Unexpected balance perturbation-training using Re-Step™ technology led to an improvement in dynamic balance and gait velocity in chronic ABI participants. The advantage of Re-Step™ technology training compared to conventional balance training should be further examined.

## Introduction

Acquired Brain Injury (ABI) i.e., Traumatic Brain Injury (TBI) and stroke is considered to be a significant and global public health issue. Each year approximately 69 million individuals worldwide suffer a TBI, and 12 million suffer a stroke (Dewan et al., 2019; Gorelick et al., 2019; Katan et al., 2018; Donkor, 2018). Both etiologies lead to dynamic balance impairments (McCulloch et al., 2010; Lord et al., 2004). Dynamic balance impairment is a risk factor for falls post-ABI, and has a significant impact on physical function, activity, community participation and quality of life years after the injury (McCulloch et al., 2010; Lord et al., 2004; Peterson et al., 2015; Klima et al., 2019; Medley et al., 2006). About 30% to 65% of TBI survivors and 73% of community-dwelling stroke survivors, report continual problems in dynamic balance control and gait (Klima et al., 2019, Forster et al., 1999) and a fall within 6 months post-stroke (Yates et al., 2002). ABI survivors demonstrate also reduced gait stability and adjustments of gait patterns when faced with environmental challenges, such as avoiding obstacles (Hyndman et al., 2006; Van Swigchem et al., 2013).

Several rehabilitation technologies have been suggested to improve dynamic balance control during gait post-ABI (Tally et al., 2017; Handelzalts et al., 2019). Tally et al. (2017) found that treadmill training improves standing balance control, but not dynamic balance during gait. Handelzalts et al., (2019) showed that unexpected perturbations exercises while treadmill walking contribute to improvement in reactive balance. However, these methods are applicable only for a clinical setting, and did not show any superiority over traditional balance training in terms of proactive balance measures (Handelzalts et al., 2019). Lastly, the impact of dynamic balance rehabilitation during the chronic stages of ABI is still not clear.

Previous studies report a decrease in training gains in TBI and stroke participants as a function of time since injury (Hayden et al., 2013; Llorens et al., 2018), and lack of improvement in sway in stroke participants that enrolled in the training program more than 2 years after injury (Llorens et al., 2018).

To address this shortage in effective rehabilitation modalities at the chronic phase post brain injury, we aimed to examine the efficacy of training using mechatronic shoes (Re-Step™ technology) which introduce unexpected perturbations during walking (Bar-Haim et al., 2013), to dynamic balance and gait velocity in chronic ABI participants. The Re-Step™ training is not limited to the clinical setting and can be performed at home and in the community with supervision of a therapist or a caregiver. We hypothesized that participants post-ABI at the chronic phase will improve their dynamic balance control and mobility following long-term unexpected balance perturbation training.

## Methods

### Trial design

Interventional prospective trial. The study was approved by the Research Ethics Committee of the Reuth Rehabilitation Medical Center, Tel-Aviv, Israel. The ClinicalTrials.gov identifier number is NCT02215590. Participants signed an informed consent, after they underwent a neurological and functional assessment at Reuth Rehabilitation Medical Center.

### Participants

Participants post-ABI (ischemic stroke or TBI). They were approached using a database of hospitalized patients in Reuth Rehabilitation Medical Center, Tel-Aviv, Israel, and by using an adds that was published in a local newspaper. Inclusion criteria were: Age range from 18-80 years; Residual dynamic balance impairment due to ABI; At least 12 months post-ABI (TBI or ischemic stroke) before recruitment; Ability to walk at least 10 meters with or without an assistive device; Drug therapy unchanged for one month prior to trial and during the entire trial period; Above 19 on the Montreal Cognitive Assessment test (MoCA) (De Guise et al., 2014; Toglia et al., 2011). Exclusion criteria were: Presence of an acute progressive neurological; systemic, or musculoskeletal disorder affecting gait and balance; Severe visual or hearing impairment; Pulmonary or cardiac condition impairing exercise endurance; Significant psychiatric disorder; Alcoholism or drug use; severe arthritis, arthralgia, fractures, or low back pain.

### Intervention protocol

**The participants performed 22** sessions, given twice a week, separated by at least 2 days. Each session began with several warm-up exercises (e.g., mobilization and strengthening) for 10 minutes, followed by perturbations intervention using the Re-Step™ for up to 40 minutes with breaks, as the participant needed. The session ended with 10 minutes of cool-down exercises, stretching and relaxation. A mechatronic shoes system (Step of Mind, Israel) developed for training and gait-rehabilitation of individuals with brain damage (Bar-Haim et al., 2013) was used to train balance during gait. The system is comprised of a software and mechatronic shoes. Each shoe has four pistons underneath its shoe that perturbed gait by sole inclinations (Figure 1) (see more details in Bar-Haim et al., 2013). During the swing phase of gait, the system introduces a range of sole inclinations: up to 6 degrees in the coronal and sagittal planes, and up to 18 mm vertical displacements (height), with predetermined maximum and minimum inclination values for defining the degree of the perturbations. Prior to the treatment session, a software determines a random sequence of vertical displacement of the pistons and the inclination levels that will be introduced during the session in terms of perturbation frequencies, defined as the number of steps between the changes of sole position. The changes in the inclination levels of the sole and the height level of the pistons are introduced during the swing phase (i.e., shoe is unloaded), and cannot be detected by the subject prior the stance phase of gait (details in Bar-Haim S et al., 2013). During the training sessions and between sessions, the vertical displacement of the pistons and the inclination levels (perturbation magnitude) and frequency of the perturbations were increased within a physically tolerable range that was assessed individually for each participant by a physical therapist (Table 1). The criterion for an increase was stepping for 20 meters without losing stability and without a need for physical assistance. The instruction to the participants was “Try to continue walking without falling”. During the perturbation session, participants were exposed to a set of walking exercises to stimulate real-life challenging situations. The exercises were: 1) Walking forward, backward, and sideways; 2) Dribbling a ball; 3) Ascending and descending stairs; 4) Stepping over obstacles; and 5) Tandem walking.

**Table 1:**
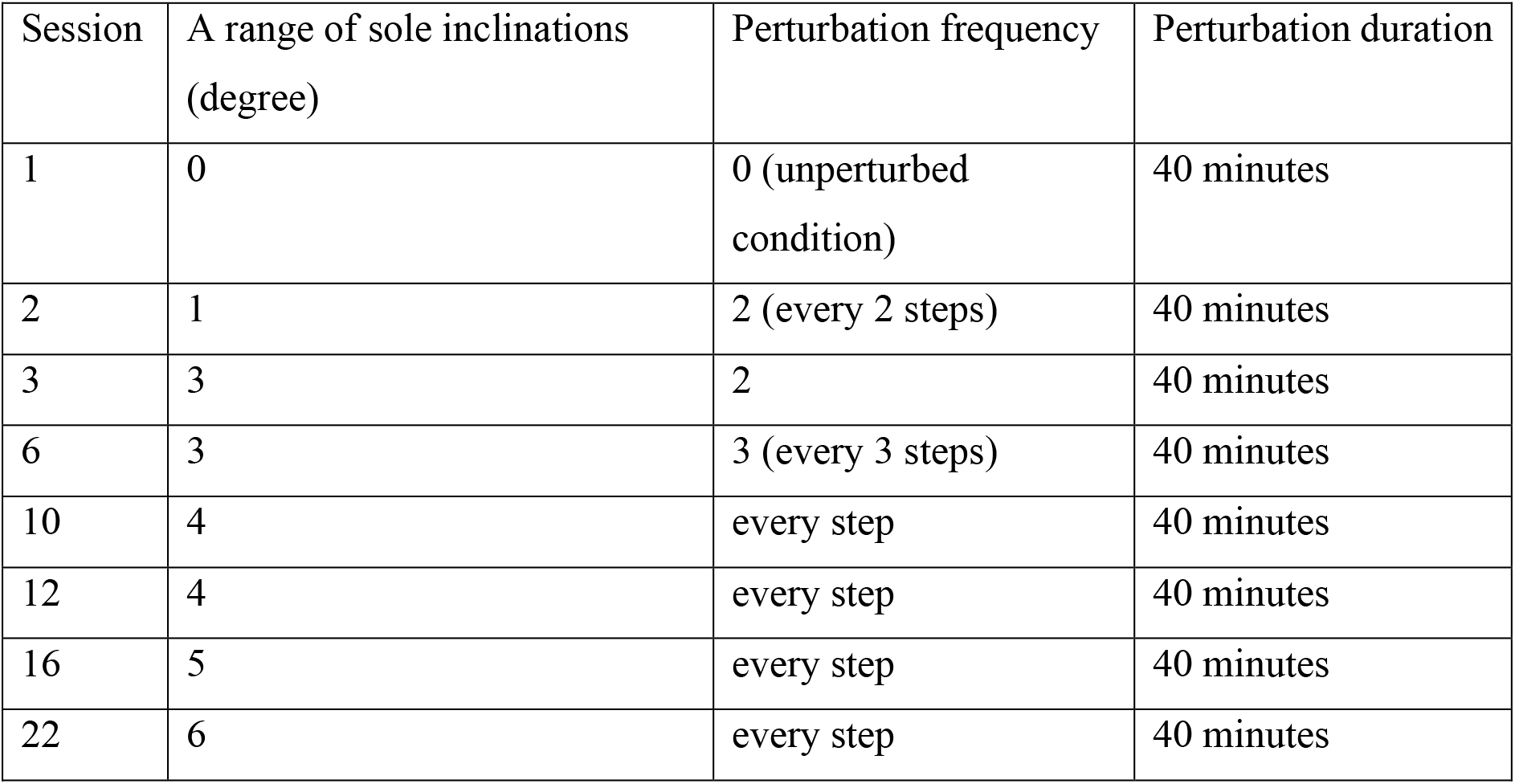
Protocol of training. (number of sessions, sole inclination levels and frequency and duration of perturbations)

**Figure 1:**
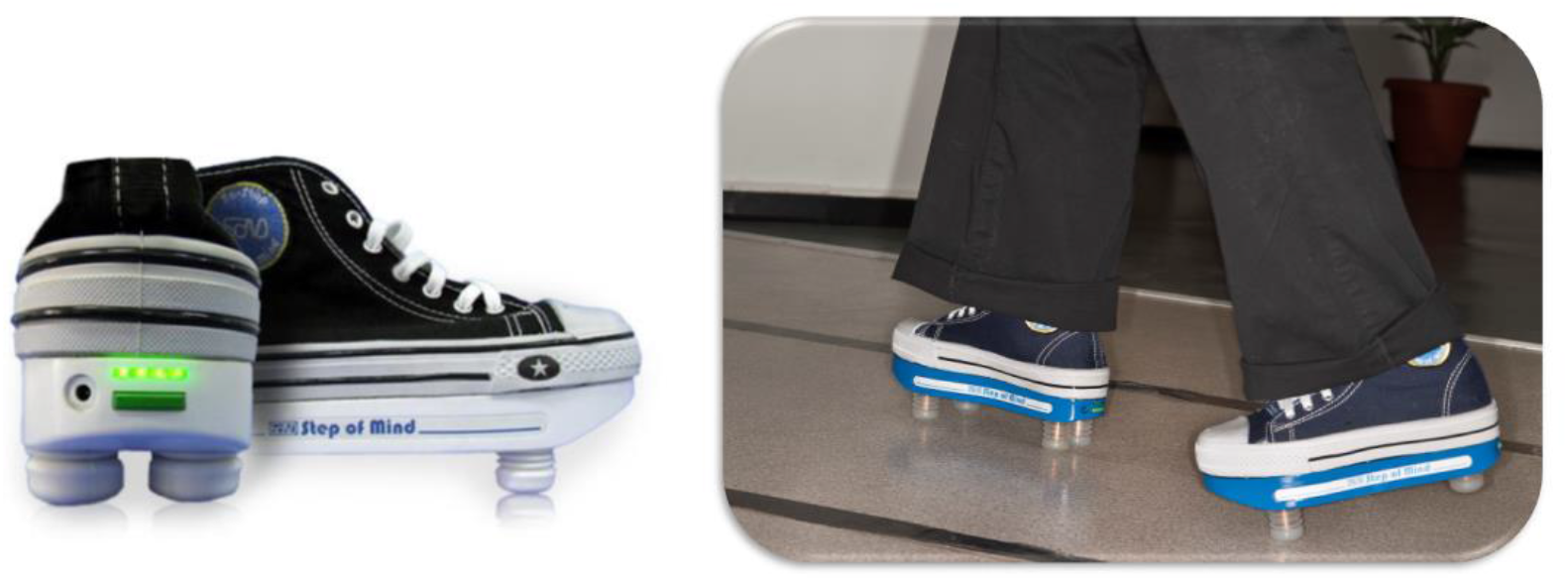
Re-Step ™ mechatronic shoes.

For safety, a physiotherapist and an assistant (physical trainer) accompanied a single participant during the Re-Step ™ training. Each session was documented in a special treatment diary by a physiotherapist. The intervention was conducted at Reuth Rehabilitation Medical Center, Tel-Aviv, Israel.

Participants were assessed pre-intervention (T_1_) and immediately post 3-month intervention (T_2_). For a sub-group of five participants post-stroke, we conducted two pre-intervention assessments (T_0_) and (T_1_), separated by one month without an intervention, before they began the intervention. Assessments were conducted by two experienced physical therapists. The interventions were administered by two additional experienced physical therapists that were blinded to the outcomes of the assessments.

### Outcome measures

#### Primary outcome measure

Community Balance and Mobility (CB&M) scale. This scale is valid and reliable for assessing difficulties in ambulation and balance skills which are needed for community integration in individuals with stroke (Knorr et al., 2010; Miller et al., 2016), and adults with traumatic brain injury (Howe et al., 2006; Inness et al., 2011). The scale includes 13 tasks requiring multitasking and complex motor tasks which are necessary to independent functioning in the community (e.g., Unilateral stance, forward to backward walking, descending stairs, crouching for picking up and object from the floor and tandem walking. See Howe et al., 2006, for full list of CB&M tasks). Higher scores indicate better balance and mobility skills (maximum possible score=96). The Minimal Detectable Change (MDC) of the CB&M scale for adults post-ABI at the chronic phase is not established yet.

#### Secondary outcome measure

10 Meter Walking Test (10MWT) which is a valid (Scrivener et al., 2014), and reliable measure for gait velocity in the individuals with stroke (Tyson et al., 2009; Collen et al., 1990). Participants were asked to walk a 14-meter track at a self-paced speed. The middle ten meters were timed. Time was measured by a handheld stopwatch (reliable as an automatic timer in measurements of gait speed) (Peters et al., 2013). Velocity was computed based on the track length and time parameters. 10MWT was added to the behavioral assessments after the begging of the experiment and was performed only on the stroke participants. The MDC for participants post-stroke at the chronic phase is equal to 0.2 m/sec (Hiengkaew et al., 2012; Lewek et al., 2011).

### Sample size estimation

The sample size was based on the CB&M outcome measure. Effect size was calculated based on the following hypothesis: H_0_=0, the H_1_=8. Effect size was based on a previous study in TBI population (MDC=8) (Tefertiller et al., 2019), and standard deviation (16) was based on another study (Inness et al., 2011). Alpha of 0.05. The result of the power analysis showed that a total sample of 27 was required to achieve a power of 0.8. The sample size that was chosen for the study was 40, taking into consideration the expected compliance with the protocol and dropouts. Power analysis was conducted using the G*Power software (version 3.1.9.6. written by Franz Faul, University of Kiel, Germany) (Faul et al., 2007).

### Statistical methods

For statistical calculations, we used SPSS statistics (SPSS for Windows, Version 16.0; SPSS Inc., Chicago). The significance level was set to *p*<0.05 for all statistical tests. Descriptive statistics was used in order to present participants’ characteristics. The Shapiro-Wilk test was used to test the assumption of normal distribution measure deviations from normality (Shapiro-Wilk test *p>0*.*05*). Paired t-test was used for within-subject analyses (pre-intervention vs. post-intervention). Wilcoxon test was used for the subgroup analysis, to compute the differences between two baseline functional level (T_1_-T_0_) and between post-intervention functional level and the first baseline functional level (T_2_-T_0_). The effect size (ES) (Cohen’s *d*) for the within subject design was calculated by dividing the mean difference between pre and post-intervention by the pooled standard deviation. We used the following guidelines for interpreting effect size magnitude: (0.01) = very small, d (0.2) = small, d (0.5) = medium, d (0.8) = large, d (1.2) = very large, and d (2.0) = huge (Sawilowsky et al., 2009).

### Sub-group analysis

To examine if the change in CB&M score is due to the intervention and not due to factors such as the re-exposer to the CB&M scale, we conducted two baseline assessments in a subgroup for five participants post-stroke, with no study-specific interventions in between (Figure 2). The time difference between the assessments was 57.4 ±10.3 days. We compared the change in CB&M score due to the exposer to the task and to the intervention (T_0_ and T_2_) with the change in CB&M score due to the exposure to the test (T_0_ and T_1_).

**Figure 2:**
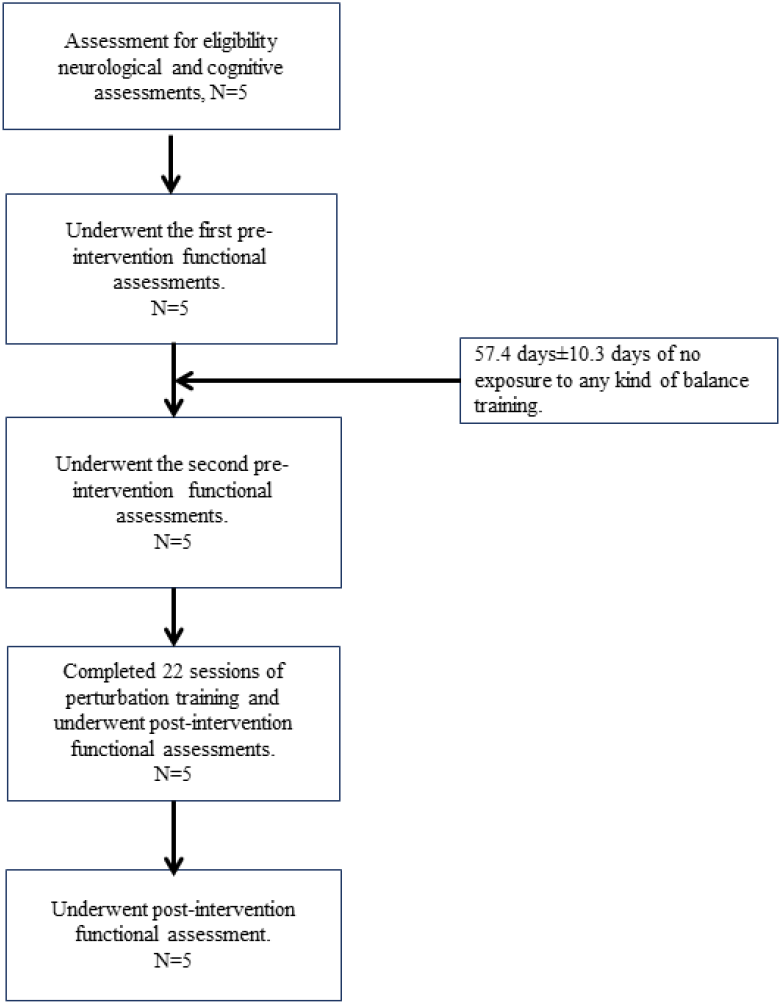
Sub-group analysis flowchart. Abbreviations: N: sample size.

## Results

### Participants’ characteristics

We recruited a convenience sample of 119 participants, all were screened by a neurologist. 41 participants were selected and underwent pre-interventional functional assessments (T_1_). Six participants dropped out due to loss of interest and transportation difficulties. 35 participants completed the 22 sessions of perturbation training and underwent post-interventional functional assessment (T_2_), figure 3 present the flow chart of this study. The time difference between the assessments was 117 days±14.14 days. Overall, the clinical analysis included the data of 35 participants (see Table 2).

**Table 2:** Baseline Characteristics of all Participants. Values are means ±SD

|  |  |
| --- | --- |
|  | N=35 |
| Age (year) | 56.94±15.28 |
| % male | 80 % |
| Height (cm) | 170.57±8.53 |
| Weight (kg) | 75.05±13.38 |
| MoCA score | 25.08±2.48 |
| Diagnosis<br>(stroke/TBI) | stroke=21<br>TBI=14 |
| Time since injury<br>(years) | 9.6±13.46 |
| Use of assistive<br>device for walking<br>(No/Yes) | 23/12 |

Abbreviations: N=sample size, cm=centimetres, kg=kilograms, TBI=traumatic brain injury

**Figure 3:**
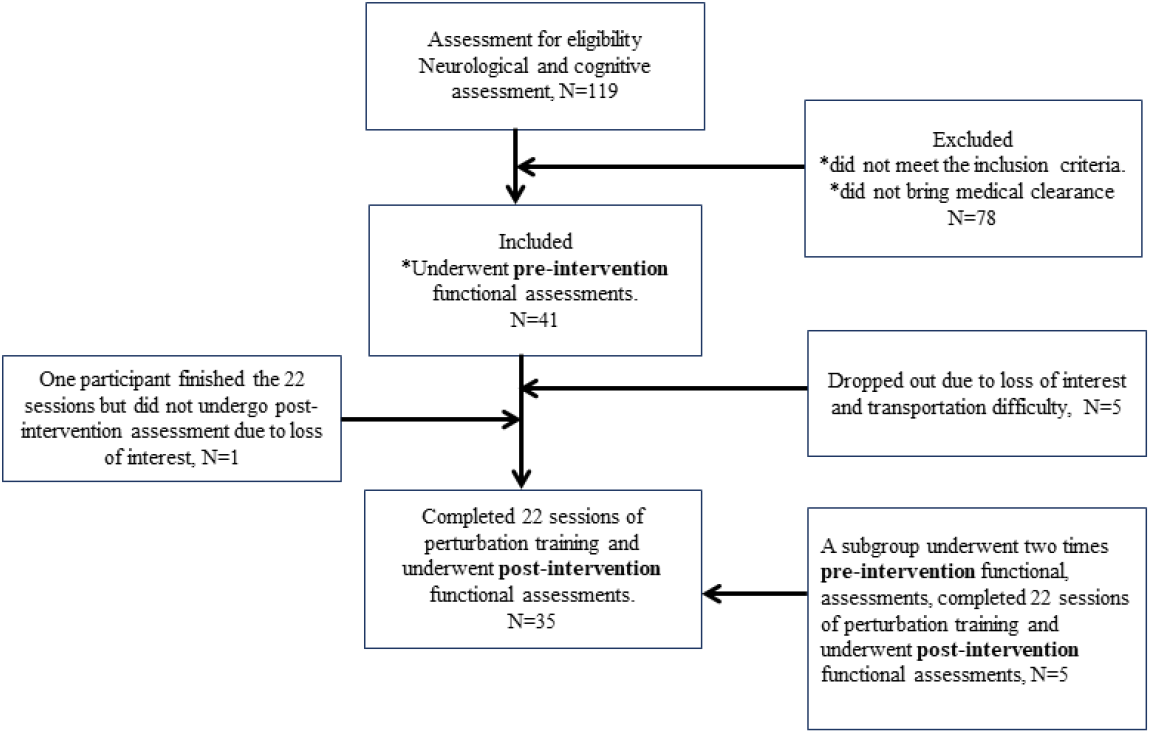
Trail flowchart. Abbreviations: N: sample size.

### Dynamic balance control

CB&M scores changed significantly from 35.87±16.2 pre-intervention (T1) to 42.89±17.04 post intervention (T2) (*p*<0.001) with a large effect size (EF=0.94) (Figure 4A). Twenty-nine (82.85%) participants showed higher scores post-intervention in comparison to the pre-intervention scores (improved), while five (14.28%) participants showed lower scores post-intervention and one (2.85%) showed no change (Figure 4B).

**Figure 4A:**
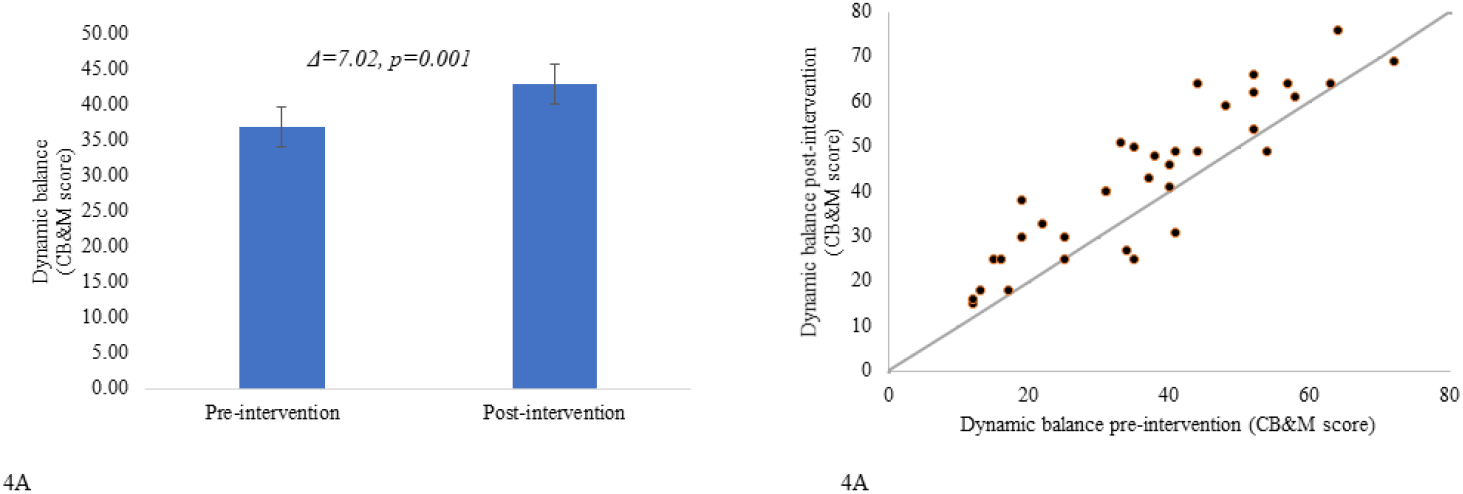
CB&M score pre- and post-intervention. The bar graph presents the CB&M score (y-axis) pre-intervention and post-intervention (x-axis). Error bars represent standard error. **Figure 4B:** A scatter plot graph presents the CB&M score distribution of the single participant (y-axis) post-intervention, (x-axis) pre-intervention (black circles). The gray line depicts the identity line (x=y).

### Gait velocity

The 10MWT scores changed significantly from 0.88 m/sec±0.34 pre-intervention (T_1_) to 0.95 m/sec ±0.32 post-intervention (T_2_) (*p*=0.02) with a medium effect size (ES=0.6) (Figure 5A the majority of participants showed an increase in gait velocity following the treatment; 16 (76.19%) participants showed higher post-intervention walking velocity in comparison to the pre-intervention velocity. four (19.04%) participants showed lower walking velocity post-intervention and one (4.76%) participant showed no change in the gait velocity post-intervention (Figure 5B).

**Figure 5A:**
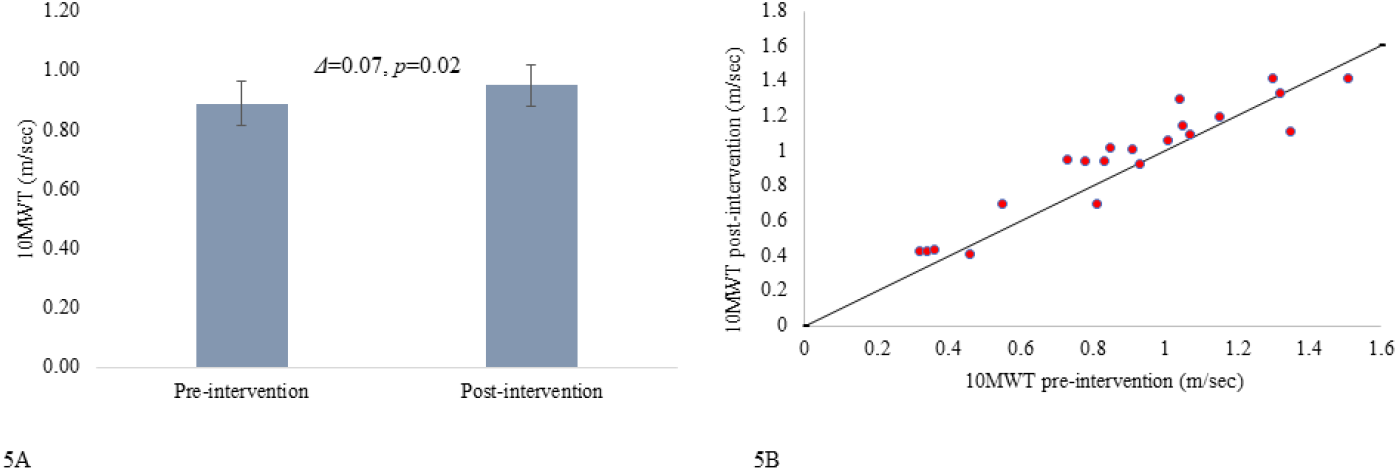
10MWT self-paced velocity (m/sec) pre-and post-intervention. A bar graph presents the 10MWT score (m/sec) (y-axis) pre-intervention and post-intervention (x-axis). Error bars represent standard error. **Figure 5B:** A scatter plot graph presents the 10MWT score distribution of the single participant (y-axis) post-intervention, (x-axis) pre-intervention (pink circles). The black line depicts the identity line (x=y).

### Sub-group analysis

We compared the change in CB&M score due to the exposer to the task and to the intervention (T_0_ and T_2_ (**Δ**_T2-T0_ CB&M=12.2±9.09)) with the change in CB&M score due to the exposure to the test (T_0_ and T_1_ (**Δ**_T1-T0_ CB&M=4.8±4.6)) (baseline characteristics of participants are listed in Table 3).

**Table 3:** Baseline Characteristics of Participants in the subgroup analysis. Values are means ±SD.

|  |  |
| --- | --- |
|  | N=5 |
| Age (year) | 68±6.2 |
| % male | 80% |
| Height (cm) | 171.8±8.01 |
| Weight (kg) | 77.4±12.4 |
| MoCA score | 26.6±2.5 |
| Diagnosis | stroke= 5 |

Abbreviations: N=sample size, cm=centimetres, Kg=kilograms

The results indicated a statistically significant difference (Z=-2.02, *p*=0.04) between both CB&M measures (Figure 6), suggesting that the training led to improvement in CB&M beyond the improvement in CB&M due to the exposure to the test itself.

**Figure 6:**
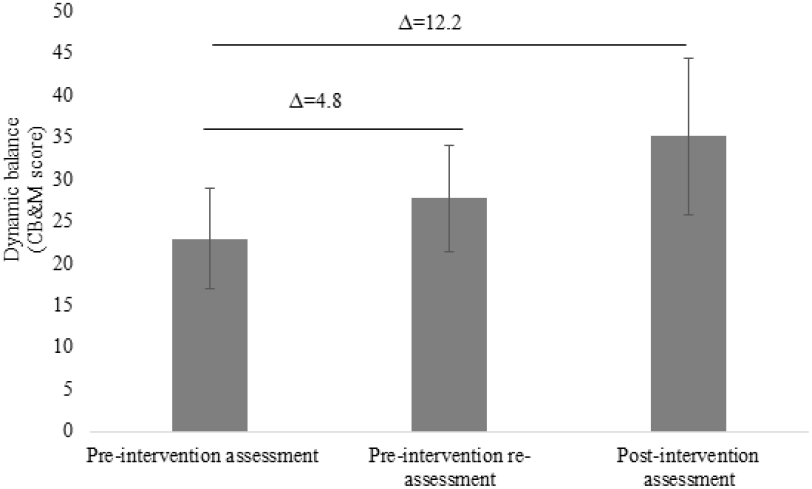
CB&M score at three time points: twice pre-intervention and once post-intervention. A bar graph presents the average CB&M score (y-axis) and three time points (x-axis): pre-intervention assessment, pre-intervention re-assessment, and post-intervention assessment. Error bars represent standard error.

For the same five subjects we conducted the sub-group analysis for the 10MWT data. The difference between **Δ**_T1-T0_ (0.09±0.08 m/sec) and **Δ**_T2-T0_ (0.15±0.14 m/sec) was not significant (*Z*=-0.94, *p*=0.34).

For all participants, there was neither any report of discomfort nor serious adverse effect during the intervention period. Four cases of losing balance and one case of crouching were reported by the therapists. These cases required the assistance of the accompanying therapist in order to prevent a fall. In all loss-of balance cases, the participants completed the training.

## Discussion

The current study is promising and supports our hypothesis that long-term unexpected balance perturbation training using Re-Step™ mechatronic shoes improves dynamic balance control in patients post-ABI at the chronic phase and self-paced gait velocity post-stroke. Participants showed a statistically significant change in dynamic balance control and gait velocity post-training as measured using the CB&M and the 10MWT despite ABI chronicity (9.6±13.46 years). The change in dynamic balance control post-intervention using Re-Step™ mechatronic shoes is in line with a previous pilot trial that showed an improvement in balance during standing and gait stability for two participants post-stroke with hemiparesis and two participants with cerebral palsy when compared to four healthy controls (Bar-Haim et al., 2013). The subgroup analysis confirmed that the change in the CB&M score post-intervention was higher than the expected effect of re-exposure to the CB&M examination.

### Clinical relevance and implication

The CB&M is an appropriate measure that evaluates gait, balance and mobility limitations of higher functioning subjects who are living in the community but may be at risk of falling (Miller et al., 2016; Howe et al., 2006; Inness et al., 2011). However, the MDC of the CB&M measure in participant post-ABI at the chronic phase is not established yet. A recent published study addressing balance deficits following TBI at the chronic phase suggested using an MDC value of 8 as a change in the CB&M score that the therapist can interpret as a true change in balance and not related to a measurement error (Tefertiller et al., 2019).

Tefertiller et al., (2019) presented an improvement in CB&M score post an in-home 12-week physical therapy (virtual reality or conventional balance training) of 7.7 and 7.8 respectively in subjects post-TBI at the chronic phase. Similarly, our participants showed CB&M score improvement post-perturbation training of Δ=7.02. When examining the proportion of participants that showed greater CB&M improvement than the suggested MDC (MDC=8), 48% of participants met or exceeded MDC in the previously mentioned study and 45.7% of the participants in our study met or exceeded the MDC (Tefertiller et al., 2019). The fact that the CB&M changes following our intervention were comparable to those seen both in the experimental and in the control group in the previously mentioned study (Tefertiller et al., 2019), suggesting that the additive contribution of the applied perturbations is limited.

The MDC of 10MWT that was reported in participants post-stroke is equal to 0.2 m/sec (Hiengkaew et al., 2012; Lewek et al., 2011). In our study the gait velocity change (**Δ**_**T2-T1**_) was lower (0.07 m/sec). in our study Only 9.5% of the participants exceeded the MDC yet the change post-intervention was statistically significant. It is accepted to classify walking ability if it is limited for household or community ambulation according to gait velocity that is measured by 10MWT (Perry et al., 1995; Bowden et al., 2008). According to the classification and cut-off scores of walking handicap in the stroke population (Perryet al., 1995), gait ability of the stroke participants in our study improved for five participants (23.8%), did not change for 15 participants (71.4%) and deteriorated for one participant (4.7%) (Table 4).

**Table 4:** number of participants, classification of gait ability (velocity m/sec), delta of gait velocity (improvement/ no change/ deterioration)

| Number of participants | Walking ability pre-intervention | Walking ability post-intervention | Improvement/no change /deterioration in gait velocity/ability |
| --- | --- | --- | --- |
| 3 | household ambulators (<0.4 m/sec) | limited community ambulators (0.4 m/se-0.8 m/sec) | Improvement in gait velocity and gait ability |
| 2 | limited community ambulators (0.4 m/se-0.8 m/sec) | community ambulators (>0.8 m/sec) | Improvement in gait velocity and gait ability |
| 1 | community ambulators (>0.8 m/sec) | limited community ambulators (0.4 m/se-0.8 m/sec) | Deterioration in gait velocity and gait ability |
| 10 | community ambulators (>0.8 m/sec) | community ambulators (>0.8 m/sec) | Improvement in gait velocity and no change in gait ability |
| 1 | community ambulators (>0.8 m/sec) | community ambulators (>0.8 m/sec) | No change in gait velocity and ability |
| 1 | limited community ambulators (0.4 m/se-0.8 m/sec) | limited community ambulators (0.4 m/se-0.8 m/sec) | Improvement in gait velocity but no change in gait ability |
| 1 | limited community ambulators (0.4 m/se-0.8 m/sec) | limited community ambulators (0.4 m/se-0.8 m/sec) | Deterioration in gait velocity but no change in gait ability |
| 2 | community<br>ambulators (0.4<br>m/se-0.8 m/sec). | community<br>ambulators (0.4<br>m/se-0.8 m/sec). | Deterioration in<br>gait velocity but<br>no change in<br>gait ability |

### Study limitations

(a) The lack of a control group that did not perform any rehabilitation treatment. Although all participants were at the chronic phase of the injury, where spontaneous recovery is unlikely (Cassidy et al., 2017), data from a control group that received no treatment is important for substantiating the effectiveness of the Re-Step™ training. (b) The lack of a long-term follow-up of the clinical assessment. Participants were assessed after the end of training, but due to the low compliance for the follow up assessment, information about the long-term retention of the training gains is missing. (c) The sample included two types of brain injury etiologies: stroke and TBI. This heterogeneity may have contributed to greater variability in the clinical results.

## Conclusion

Our findings indicate that rehabilitation of dynamic balance at the chronic phase post-ABI is possible and time since injury does not limit rehabilitation utility when using Re-step™ technology. Using perturbation-based balance training, such as the Re-step™ technology may be an effective intervention for improving dynamic balance control post-ABI at the chronic phase at the clinic or at home. Our results should be followed by future studies for evaluating the superiority of our intervention with respect to conventional balance training.

## Data Availability

the data is available

## Source of Funding and acknowledgments

The Israel Insurance Association (R.A.), Association of Life Insurance Companies of Israel LTD. The funders had no role in study design, data collection and analysis, decision to publish, or preparation of the manuscript.

## Abbreviations

ABI: Acquired Brain Injury
CB&M: Community Balance and Mobility
MDC: Minimal Detectable Change
MoCA: Montreal Cognitive Assessment
TBI: Traumatic Brain Injury
10MWT: 10 Meter Walking Test

